# Unveiling the Longitudinal Journey: Three-Year Follow-up of Women with MINOCA and INOCA in a Specialized Heart Centre

**DOI:** 10.1101/2024.06.24.24309437

**Authors:** Emilie T. Théberge, Elizabeth Burden, Katrina Leung, Mahraz Parvand, Natasha Prodan-Bhalla, Karin H. Humphries, Tara L. Sedlak

## Abstract

**Background:** Myocardial infarction with no obstructive coronary arteries (MINOCA) and ischemia with no obstructive coronary arteries (INOCA), are female predominant conditions, with a lack of clinical trials guiding medical management for the common underlying vasomotor etiologies. Long-term outcomes of (M)INOCA patients following attendance at a women’s heart center (WHC) are lacking.

**Methods:** Women diagnosed with MINOCA (n=51) or INOCA (n=112) were prospectively followed for 3 years at the Vancouver WHC. Baseline characteristics, diagnoses, chest pain type, major adverse cardiac events, hospital encounters, medications, and Seattle Angina Questionnaire (SAQ) responses were compared between baseline and 3 years. Chi-squared tests were used to compare categorical variables, with Students’ t-tests for continuous variables.

**Results:** MINOCA patients had significantly more non-exertional chest pain and diagnoses of vasospasm than INOCA patients, who had more exertional chest pain and diagnoses of coronary microvascular dysfunction. Following baseline, both groups had significant reductions in cardiovascular emergency room visits, with INOCA patients also experiencing fewer cardiovascular hospitalizations. At 3 years, the most commonly prescribed medications were calcium channel blockers, long-acting nitrates and beta blockers, with MINOCA having more acetylsalicylic acid and INOCA more short-acting nitrates and ranolazine prescriptions. Both groups observed significant improvements in SAQ scores, with greater improvements observed in INOCA patients. Patients with depression or prescribed ranolazine at 3 years had worse SAQ scores at baseline.

**Conclusions:** Three-year outcomes of (M)INOCA patients indicate that the WHC’s comprehensive care model effectively improves diagnostic clarity, reduces hospital encounters, optimizes medication management, and improves self-reported patient well-being.

## Introduction

Nonobstructive coronary syndromes, including myocardial infarction with no obstructive coronary arteries (MINOCA) and ischemia with no obstructive coronary arteries (INOCA), present unique challenges in cardiology as a result of their enigmatic nature and clinical implications. Both are characterized by less than 50% stenosis in any major coronary artery, MINOCA in the setting of myocardial infarction (MI) and INOCA with ischemia. MINOCA afflicts up to 6% of individuals experiencing an acute coronary syndrome, more so if they are women.^1,2^ Similarly, two thirds of angiograms for women with suspected cardiac ischemia reveal INOCA, at a prevalence twice that seen in men. Predominant underlying ischemic etiologies of these two syndromes (“(M)INOCA”) include coronary vasospasm and coronary microvascular dysfunction (CMD), conditions that are more common in women and are found to have more nontraditional risk factors as compared to those with obstructive coronary artery disease (CAD).^3^

While patients with (M)INOCA were previously considered benign and managed conservatively, these entities have gained attention in recent years due to their association with adverse cardiac outcomes and mortality.^4,5^ A 2022 systematic review and meta-analysis assessing sex-related discrepancies in MINOCA outcomes revealed a higher incidence of major adverse cardiac events (MACE) in women as compared to men within 24 months, largely driven by a greater incidence of stroke.^6^ This holds true in INOCA patients, with a 2.55-fold higher risk of MACE in women with INOCA compared to women with no CAD within one year following angiographic investigations.^7^ Beyond this, patients with nonobstructive coronary syndromes are burdened with poor physical functioning, recurrent angina, and reduced quality of life.^8,9^

At this juncture, there is a lack of randomized controlled trial data to provide guidance on medical treatment strategies in patients with (M)INOCA. The recent clinical practice update from the Canadian Cardiovascular Society and Canadian Women’s Heart Health Alliance acknowledges this and how, as such, prescribing approaches are on the basis of expert opinion and a personalized strategy to the underlying etiology.^10^ This is supported by the CorMicA trial, which demonstrated that stratified medical therapy based on invasively diagnosed etiology significantly improved treatment satisfaction, angina severity, and quality of life.^11^

Women’s Heart Centres (WHC) provide specialized and comprehensive care tailored specifically to the unique needs of women.^12^ There is growing recognition of specialized WHCs as a sustainable solution to address the disparities faced in this understudied population, particularly given the ongoing confusion with regard to standardized terminology and diagnostic criteria.^13^ In our previous study, Parvand et al. showcased 1- year prospective data suggesting WHC enrolment resulted in higher diagnostic yield, improved risk-factor management, and reduced angina hospitalization in women presenting with (M)INOCA.^14^ Furthermore, due to the heterogeneity of the (M)INOCA patient population, access to a WHC may allow for tailored prescribing practices.

Overall, there remains a paucity of data evaluating the role of specialized WHCs in the longitudinal follow-up of (M)INOCA patients. The aim of this work was to provide more detailed insights into the outcomes, diagnoses, longer-term therapeutic management, and modifiers of patient-reported outcomes for this population after 3-years of follow-up in a WHC.

## Methods

### Women’s Heart Centre Cohort

Female patients with MINOCA or INOCA referred to the Leslie Diamond WHC in Vancouver, Canada, were identified for WHC Registry enrolment as approved by the University of British Columbia’s clinical research ethics board (application #H13-03322). Patients were consented into the WHC Registry since 2015 following their baseline WHC consultation, with prospective data obtained thereafter for at minimum 3 years.

Patients were followed by a multidisciplinary team consisting of a cardiologist with specialized training in women’s heart health, a psychiatrist, a dietician, and two part-time nurse practitioners. They had access to specialized diagnostic techniques, including invasive coronary reactivity testing (CRT), adenosine stress cardiac magnetic resonance imaging (MRI) and optical coherence tomography (OCT) during angiography. Patients were seen at least once annually for regular clinical follow-up, with increased frequency as needed.

### Data Collection

Patient data obtained from questionnaires were stored using a REDCap database. Demographic, clinical and female-specific variables were captured from the baseline WHC clinical consultation and/or verbal self-report to the study coordinator and entered into the REDCap database.

### Study Outcomes

#### Baseline Characteristics

Frequency of variables captured at baseline were compared between MINOCA and INOCA: age, race(s)/ethnicity(ies), partnered status, education, working status, family history of premature heart disease, prior stroke or transient ischemic attack (TIA), dyslipidemia, diabetes mellitus, hypertension, obesity, smoking history, arrhythmia, depression, anxiety, thyroid dysfunction, migraine, Raynaud’s syndrome, chronic obstructive pulmonary disease, autoimmune disease, menopause status, number of children born, preeclampsia or gestational hypertension during pregnancy, and polycystic ovarian syndrome. Full definitions of each variable are described in the Supplemental Methods.

A key exposure variable was a history of depression and was considered present if any of the following 3 criteria were met: 1. Recent or current clinical depression diagnosis indicated in baseline consultation; 2. Baseline Patient Health Questionnaire (PHQ-9) score of ≥10 points;^15^ or 3. Currently taking (an) antidepressant(s) for depressive symptoms as confirmed by the patient self-report and/or consult notes (including referral documentation prior to WHC). This definition was used to account for self-report bias and/or inconsistent diagnostic appraisal possible between health providers.^16^ Expanding the criteria for defining depression to include any combination of these variables (antidepressant use, questionnaire response and self-report data) have been previously shown to improve validity of depression studies.^17^

#### Cardiac Diagnoses

Referral letters from primary care or specialist physicians were reviewed if the patient had undiagnosed chest pain attributed to (M)INOCA. Prior to baseline, the patients were appraised to either: a) Lack a specialized diagnosis, with referral to the WHC for diagnostic clarity, or b) Possess diagnostic evidence of an etiology underlying the (M)INOCA, with request for further medical opinion in symptom management. The (M)INOCA etiologies were classified as either a vasomotor etiology (CMD or coronary vasospasm) or “other” non-vasomotor etiology (i.e., spontaneous coronary artery dissection (SCAD), Takotsubo cardiomyopathy, myocarditis, missed plaque rupture).

Probable or confirmed CMD and coronary vasospasm were defined using the Coronary Vasomotion Disorder International Study (COVADIS) group definitions^18^ and are further described in the Supplemental Methods. The resulting diagnosis from cardiac investigations and/or medications prescribed during attendance at the WHC were identified from physician consult notes between baseline attendance and 3 years of follow-up. If patients were ascertained to have SCAD alone prior to baseline WHC attendance, they were triaged to a dedicated SCAD clinic at our centre and were not included. If patients had overlapping diagnoses of SCAD and an additional (M)INOCA etiology, they were included in the study. Similarly, if SCAD was identified following enrolment in the WHC Registry, they were included.

#### Clinical Presentation

Baseline WHC physician consult notes were reviewed for descriptors of chest pain on exertion, chest pain at rest, and/or chest pain during stress or emotional triggers. These terms were explicitly stated as such in the consult notes, and any categorization ambiguities were clarified before categorization.

Cardiac testing modalities used for ischemic, structural and overall assessment were quantified: coronary angiography (CA) or computed tomography coronary angiography (CTCA), degree of stenosis from CA/CTCA, results from CRT with adenosine, acetylcholine or ergonovine provocation during CA, exercise stress tests (ESTs), cardiac MRI with and without adenosine, myocardial perfusion imaging, and echocardiography. For the detection of microvascular dysfunction, modalities were limited to invasive CRT and stress cardiac MRI, as doppler ultrasound and PET scanning techniques are not available in the province of BC.

#### Hospital Encounters and MACE

Hospital encounters, defined as emergency room (ER) visits for chest pain, hospitalizations for any cardiovascular cause, and MACE, were quantified within the 3 years prior to the baseline WHC visit and up to 3 years post-baseline. Only electronic health record encounters in British Columbia were accessible. MACE comprised MI, stroke, heart failure, or cardiovascular death. TIAs were also separately evaluated, as they are known precursors to stroke within 1 year.^19^ For MINOCA patients, their first (index) MI was excluded from the pre-baseline hospitalization count.

#### Therapeutic Management

Medications were quantified at baseline and 3-years. The baseline medications reflect what the patient was taking prior to the WHC appointment. The 3-year medications were identified from the most recent physician consult notes, reflecting changes done in the WHC. For patients with medications data at both baseline and 3-years, changes in medications were analyzed on a per-individual basis.

#### Patient-Reported Outcomes

The Seattle Angina Questionnaire (SAQ) is a questionnaire validated in patients with cardiac chest pain that measures four dimensions of self-reported symptoms: physical limitation, angina frequency, quality of life, and treatment satisfaction.^20^ Scores were scaled to a 100-point scale prior to analysis. Scaled scores between 0-24 represent severe angina, 25-74 as moderate angina, are moderate angina, and over 75 as mild to no angina as corresponding to clinically relevant CCS angina scales.^21^ The SAQ was administered at baseline and 3 years, and changes in scores assessed by changes on the 100-point scale. The minimal clinically important difference (MCID) for the SAQ is 10 points on the 100-point scale.^20^

SAQ subdomain scores were further stratified and analyzed by the presence of depression at baseline or prescription of ranolazine by year 3. We hypothesized that patients with depression at baseline would have worse SAQ scores, as depression has been demonstrated to accentuate perceptions of pain.^22^ Further, as ranolazine is used as a second- or third-line anti-anginal agent, we hypothesized that patients prescribed this medication would have worse SAQ scores at baseline owing to more challenges to manage angina.

### Statistical Analyses

Baseline characteristics are reported as means and standard deviations (SD) for continuous variables and counts and percentages for categorical variables.

Comparisons between MINOCA and INOCA groups were conducted using Chi-squared tests or Student’s t-test for categorical and continuous variables, respectively. Statistical significance in the number of ER visits, hospitalizations and MACE events between baseline and year 3 timepoints were calculated using McNemar’s test.

Overall SAQ scores by subdomain were compared using Student’s t-tests. Two variables were investigated as modifiers of SAQ scores at baseline, at 3 years, and the effect on the difference between baseline and 3 years: 1) Depression status at baseline (yes/no) and 2) Ranolazine prescription status at 3 years (yes/no). For each analysis, Students’ t-tests were used to compare mean questionnaire scores for each modifier variable. Baseline and year 3 scores within groups were compared using paired Wilcoxon rank sum tests for each subdomain, as data were non-normally distributed. To assess if depression and ranolazine impacted the change in questionnaire scores over time, differences between baseline and 3-year scores were first calculated for each patient, then the means of each variable, stratified by the presence/absence of each modifier variable, were compared using the Student’s t-test. Linear regression models were plotted to display trends.

## Results

### Baseline Characteristics

There were 265 patients recruited between 2015 and 2020 who were considered for analysis. Excluded patients consisted of 32 patients who had withdrawn or were lost to follow-up (12%), 64 who were subsequently diagnosed with a non-(M)INOCA etiology (24%), and 6 with incomplete surveys (2%). This resulted in 163 patients (62%) eligible for analysis with complete baseline and 3-year questionnaires, of whom 51 had MINOCA and 112 had INOCA. Among demographic, clinical and female-specific variables (Table 1), there were no significant differences observed between MINOCA and INOCA patients, except for migraines, which were significantly more prevalent in MINOCA patients (23, 45%) than INOCA patients (28, 25%).

**Table 1:**
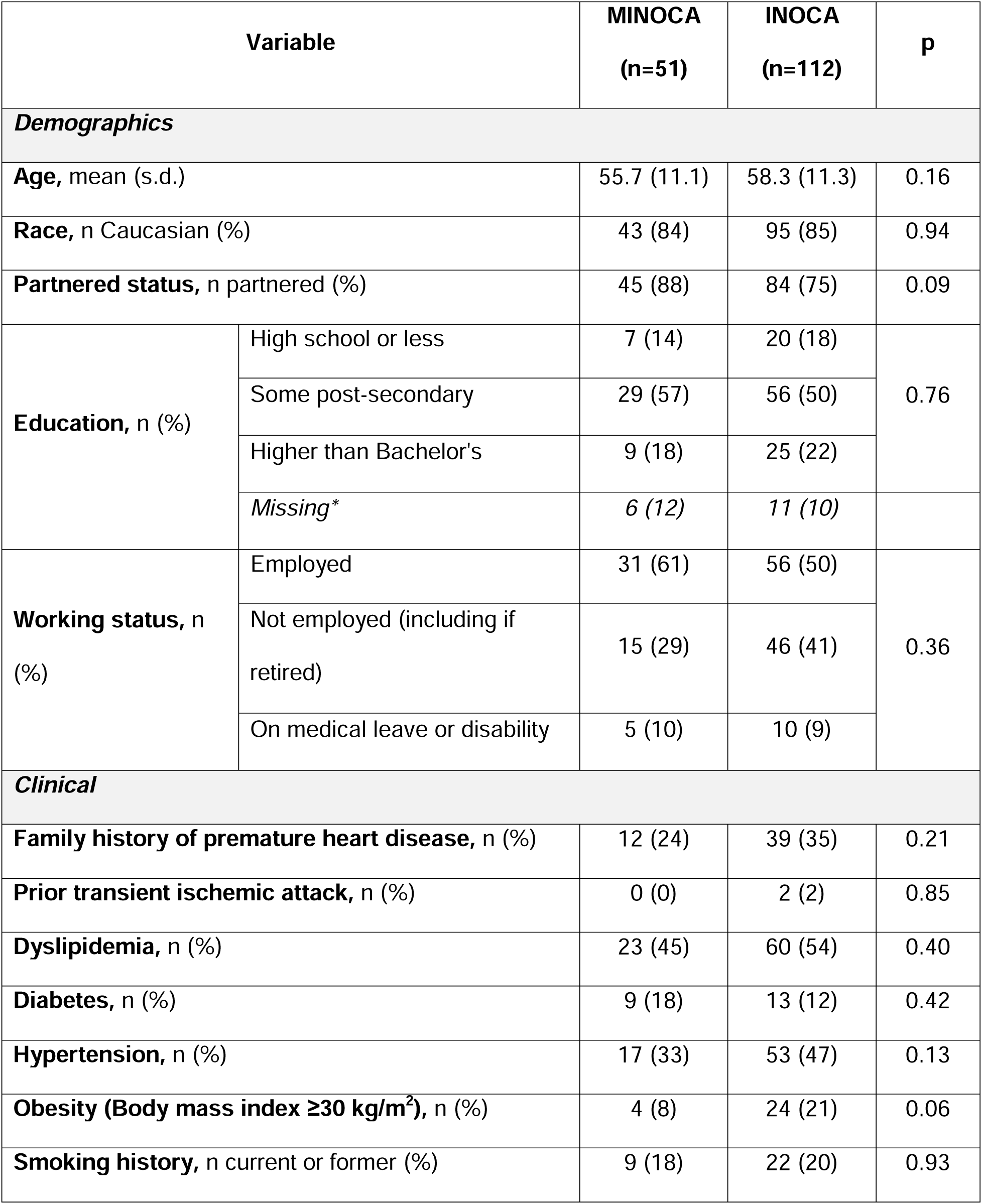
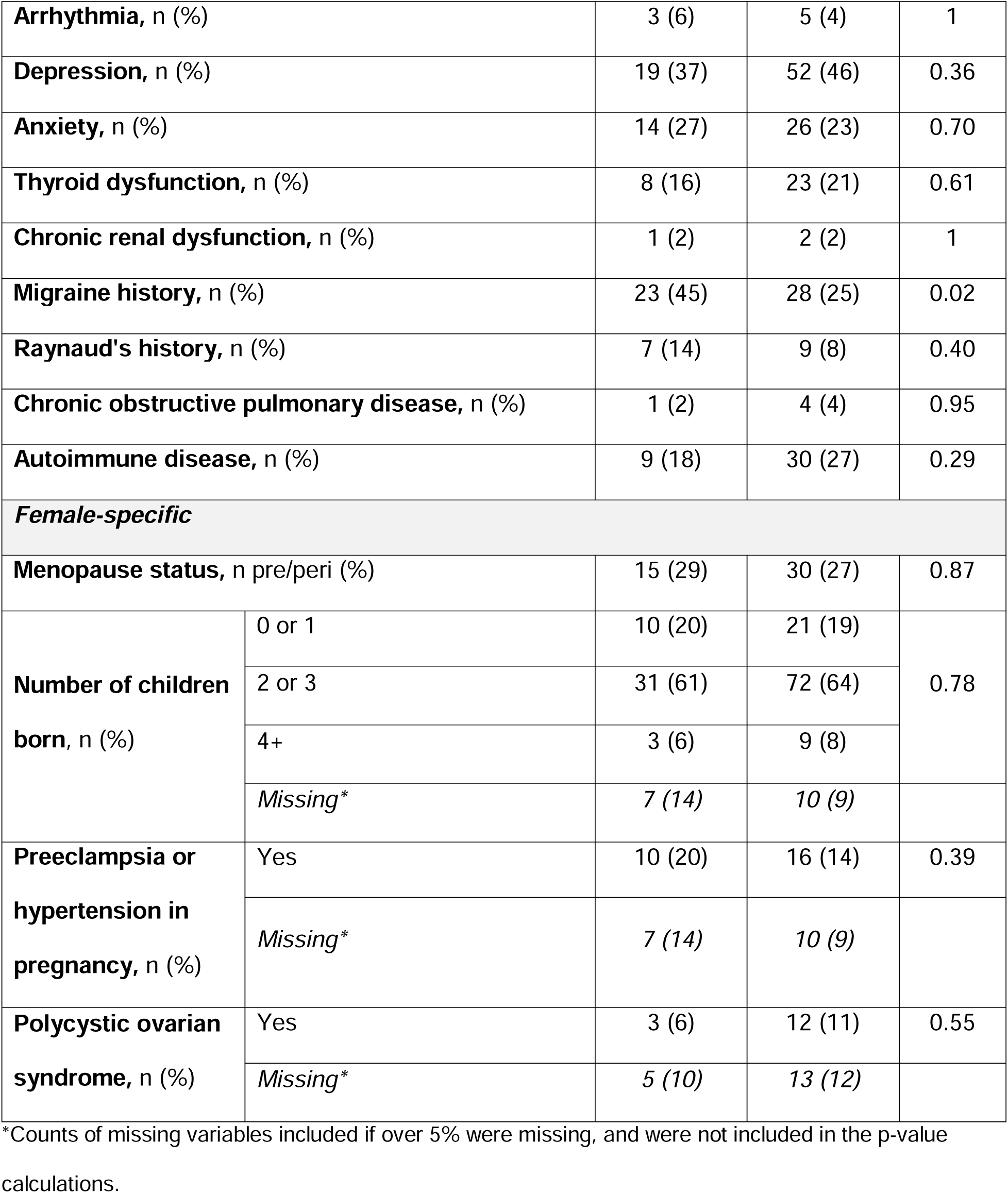
Baseline characteristics stratified by MINOCA and INOCA.

| Variable |  | MINOCA<br>(n=51) | INOCA<br>(n=112) | p |
| --- | --- | --- | --- | --- |
| Demographics |  |  |  |  |
| Age, mean (s.d.) |  | 55.7 (11.1) | 58.3 (11.3) | 0.16 |
| Race, n Caucasian (%) |  | 43 (84) | 95 (85) | 0.94 |
| Partnered status, n partnered (%) |  | 45 (88) | 84 (75) | 0.09 |
| Education, n (%) | High school or less | 7 (14) | 20 (18) | 0.76 |
|  | Some post-secondary | 29 (57) | 56 (50) |  |
|  | Higher than Bachelor's | 9 (18) | 25 (22) |  |
|  | Missing* | 6 (12) | 11 (10) |  |
| Working status, n (%) | Employed | 31 (61) | 56 (50) | 0.36 |
|  | Not employed (including if retired) | 15 (29) | 46 (41) |  |
|  | On medical leave or disability | 5 (10) | 10 (9) |  |
| Clinical |  |  |  |  |
| Family history of premature heart disease, n (%) |  | 12 (24) | 39 (35) | 0.21 |
| Prior transient ischemic attack, n (%) |  | 0 (0) | 2 (2) | 0.85 |
| Dyslipidemia, n (%) |  | 23 (45) | 60 (54) | 0.40 |
| Diabetes, n (%) |  | 9 (18) | 13 (12) | 0.42 |
| Hypertension, n (%) |  | 17 (33) | 53 (47) | 0.13 |
| Obesity (Body mass index ≥30 kg/m<sup>2</sup>), n (%) |  | 4 (8) | 24 (21) | 0.06 |
| Smoking history, n current or former (%) |  | 9 (18) | 22 (20) | 0.93 |
| **Arrhythmia, n (%)** |  | 3 (6) | 5 (4) | 1 |
| **Depression, n (%)** |  | 19 (37) | 52 (46) | 0.36 |
| **Anxiety, n (%)** |  | 14 (27) | 26 (23) | 0.70 |
| **Thyroid dysfunction, n (%)** |  | 8 (16) | 23 (21) | 0.61 |
| **Chronic renal dysfunction, n (%)** |  | 1 (2) | 2 (2) | 1 |
| **Migraine history, n (%)** |  | 23 (45) | 28 (25) | 0.02 |
| **Raynaud's history, n (%)** |  | 7 (14) | 9 (8) | 0.40 |
| **Chronic obstructive pulmonary disease, n (%)** |  | 1 (2) | 4 (4) | 0.95 |
| **Autoimmune disease, n (%)** |  | 9 (18) | 30 (27) | 0.29 |
| ***Female-specific*** |  |  |  |  |
| **Menopause status, n pre/peri (%)** |  | 15 (29) | 30 (27) | 0.87 |
| **Number of children born, n (%)** | 0 or 1 | 10 (20) | 21 (19) | 0.78 |
|  | 2 or 3 | 31 (61) | 72 (64) |  |
|  | 4+ | 3 (6) | 9 (8) |  |
|  | *Missing\** | 7 (14) | 10 (9) |  |
| **Preeclampsia or hypertension in pregnancy, n (%)** | Yes | 10 (20) | 16 (14) | 0.39 |
|  | *Missing\** | 7 (14) | 10 (9) |  |
| **Polycystic ovarian syndrome, n (%)** | Yes | 3 (6) | 12 (11) | 0.55 |
|  | *Missing\** | 5 (10) | 13 (12) |  |

\*Counts of missing variables included if over 5% were missing, and were not included in the p-value calculations.

### Cardiac Diagnoses

Most patients who were referred to the WHC with chest pain did not have a specific etiology diagnosed at entry (131, 80%). A minority (32, 20%) of the referrals requested medical management consultation for chest pain with prior diagnosed CMD (8, 5%), vasospasm (9, 6%) or other diagnoses (15, 9%).

Figure 1 demonstrates pre- versus post-WHC diagnostic changes and Table 2 summarizes the specific etiologies diagnosed after 3 years of WHC follow-up. Most had a final vasomotor diagnosis of either CMD (92, 56%) or vasospasm (42, 26%). The majority of INOCA patients had CMD (85, 76%) followed by vasospasm (18, 16%), and the majority of MINOCA patients had vasospasm (24, 47%) followed by a non-vasomotor etiology (18, 35%). There were significantly more MINOCA patients with vasospasm (47%) or another non-vasomotor etiology (35%) compared to INOCA, with 16% and 4% respectively. In contrast, significantly more INOCA patients had CMD (76%) than MINOCA patients (14%). Among the 22 patients with “other” non-vasomotor diagnoses following WHC assessment, 5 had Takotsubo cardiomyopathy only, 1 had Takotsubo with CMD, 2 had Takotsubo with spontaneous coronary artery dissection (SCAD), 5 had SCAD and CMD, 1 had SCAD and vasospasm, 1 had SCAD only, 4 had myocarditis, 1 had coronary embolism, 1 had both vasospasm and CMD, and 1 had mild plaque with aortic stenosis.

**Figure 1:**
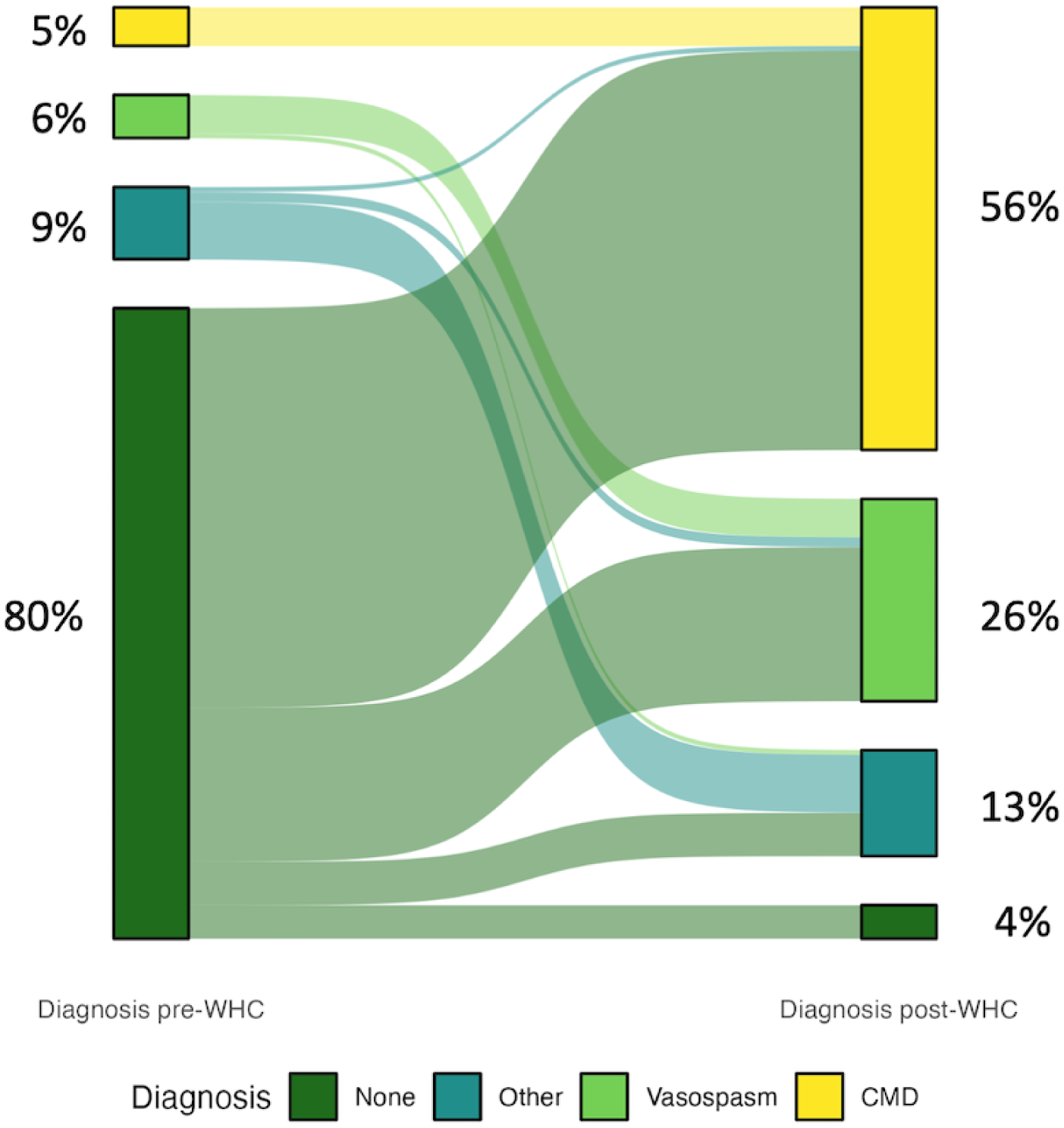
Diagnoses of 163 patients studied before and 3 years after attendance at the WHC.

**Table 2:** Comparing frequency of etiologies diagnosed following attendance at WHC.

| **Etiology** | **Total (n=163)** | **MINOCA (n=51)** | **INOCA** | **p** |
| --- | --- | --- | --- | --- |

|  |  |  | (n=112) |  |
| --- | --- | --- | --- | --- |
| Vasospasm | 42 (26) | 24 (47) | 18 (16) | < 0.001 |
| *Definitive Vasospasm* | 8 (5) | 3 (6) | 5 (4) | - |
| *Probable Vasospasm* | 34 (21) | 21 (41) | 13 (12) | - |
| CMD | 92 (57) | 7 (14) | 85 (76) | < 0.001 |
| *Definite CMD* | 14 (9) | 2 (4) | 12 (11) | - |
| *Probable CMD* | 78 (48) | 5 (10) | 73 (65) | - |
| Other non-vasomotor etiology | 22 (13) | 18 (35) | 4 (4) | < 0.001 |
| Not yet diagnosed | 7 (4) | 2 (4) | 5 (4) | 1 |

All values presented as n (%).

### Clinical Presentation

There were significant differences in the frequency of the types of chest pain observed between MINOCA and INOCA patients, with the majority of both groups displaying multiple types (Table 3, p<0.001). MINOCA patients had more presentations of chest pain only at rest (20% versus 4% in INOCA) and INOCA patients had more chest pain only on exertion (36% versus 10% in MINOCA).

**Table 3:** Chest pain type frequency by MINOCA versus INOCA groups.

| Chest pain component | MINOCA<br>(n=51) | INOCA<br>(n=112) | p |
| --- | --- | --- | --- |
| Combination of more than one chest pain type | 34 (67) | 66 (59) | <0.001 |
| Exertional only | 5 (10) | 40 (36) |  |
| Non-exertional only (at rest, sleep) | 10 (20) | 5 (4) |  |
| With stress/emotional triggers only | 2 (4) | 1 (1) |  |

All values presented as n (%).

Within a subset of patients stratified by vasospasm or CMD etiology instead of (M)INOCA, significantly more patients with CMD had an exertional component to their chest pain (80, 87%) compared to vasospasm (19, 45%) patients (Table 4, p<0.001). Conversely, significantly more patients with vasospasm had a component of non-exertional chest pain (28, 67%) compared to patients with CMD (34, 37%). The frequency of stress or emotional trigger-induced chest pain did not vary between groups.

**Table 4:** Chest pain type frequency by CMD versus vasospasm diagnosis.

| Chest pain component | CMD<br>(n=92) | Vasospasm<br>(n=42) | p |
| --- | --- | --- | --- |
| Exertional | 80 (87) | 19 (45) | < 0.001 |
| Non-exertional (at rest, sleep) | 34 (37) | 28 (67) | 0.003 |
| With stress/emotional triggers | 24 (26) | 11 (26) | 1 |

All values presented as n (%). Note chest pain totals do not add up to 100% for each diagnosis as some patients reported more than one type of chest pain.

Table 5 displays the rates of diagnostic tests for anatomic atherosclerosis and ischemia. Most patients demonstrated normal coronary arteries (MINOCA 70.6%, INOCA 60.7%) compared to those with minimal (1-24% occlusion) or mild (25-49% occlusion) CAD (MINOCA 29.4%, INOCA 33.1%). There were significantly higher rates of modalities positive for ischemia in INOCA patients compared to MINOCA, specifically EST (INOCA 38.4% compared to MINOCA 5.9%, p<0.001) and myocardial perfusion (INOCA 7.1% compared to MINOCA 0%, p=0.006). Wall motion abnormalities detected from echocardiography were significantly more common in MINOCA patients (hypokinesis 17.6% to INOCA 2.7% and akinesis 3.9% to INOCA 0%, p<0.001).

**Table 5:** Cardiac investigations for ischemia and/or structural abnormalities in patients with undiagnosed MINOCA or INOCA.

| Testing modality |  | MINOCA<br>(n=51) | INOCA<br>(n=112) | p |
| --- | --- | --- | --- | --- |
| Microvascular<br>dysfunction provocation<br>test with adenosine (CFR<br><2.5 and/or IMR ≥25) | Positive | 2 (3.9) | 12 (10.7) | 0.20 |
|  | Negative | 4 (7.8) | 4 (3.6) |  |
|  | *Not done* | 45 (88.2) | 96 (85.7) |  |
| Epicardial vasospasm<br>provocation test with<br>ergonovine or<br>acetylcholine | Positive | 3 (5.9) | 5 (4.5) | 0.59 |
|  | Negative | 0 | 2 (1.8) |  |
|  | *Not done* | 48 (94.1) | 105 (93.8) |  |
| Exercise stress test | Positive for ischemia | 3 (5.9) | 43 (38.4) | < 0.001 |
|  | Negative | 24 (47.1) | 54 (48.2) |  |
|  | *Not done* | 24 (47.1) | 15 (13.4) |  |
| Cardiac MRI | Positive for ischemia | 3 (5.9) | 6 (5.4) | 0.002 |
|  | Negative | 24 (47.1) | 23 (20.5) |  |
|  | *Not done* | 24 (47.1) | 83 (74.1) |  |
| Myocardial perfusion<br>imaging | Positive for ischemia | 0 | 8 (7.1) | 0.006 |
|  | Negative | 6 (11.8) | 31 (27.7) |  |
|  | *Not done* | 45 (88.2) | 73 (65.2) |  |
| Highest stenosis from most recent CA/CTCA prior to baseline WHC appointment | Normal | 36 (70.6) | 68 (60.7) | 0.13 |
|  | Minimal (1-24%) | 10 (19.6) | 17 (15.2) |  |
|  | Mild (25-49%) | 5 (9.8) | 20 (17.9) |  |
|  | *Not done* | 0 | 7 (6.3) |  |
| Echocardiogram wall motion findings | Normal | 29 (56.9) | 60 (53.6) | < 0.001 |
|  | Akinesis | 2 (3.9) | 0 |  |
|  | Dyskinesis | 0 | 1 (0.9) |  |
|  | Hypokinesis | 9 (17.6) | 3 (2.7) |  |
|  | Not assessed | 1 (2.0) | 0 |  |
|  | *Not done* | 10 (19.6) | 48 (42.9) |  |

All values presented as n (%).
**Acronyms:** **CA**=Coronary angiogram; **CFR**=Coronary flow reserve; **CMD**=Coronary microvascular dysfunction; **MRI**=Magnetic resonance imaging; **CTCA**=Computed tomography coronary angiogram; **IMR**=Index of microvascular resistance

### Hospital Encounters and MACE

Significantly fewer ER visits for chest pain were observed following WHC attendance for both MINOCA (92% to 51%) and INOCA patients (50% to 32%) (Table 6). INOCA patients had significantly fewer hospitalizations for any cardiovascular reason post- WHC (20% to 5%) but not MINOCA patients (4% to 8%), although for MINOCA patients, their first (index) MI hospitalization was excluded from the pre-baseline hospitalization count.

**Table 6:**
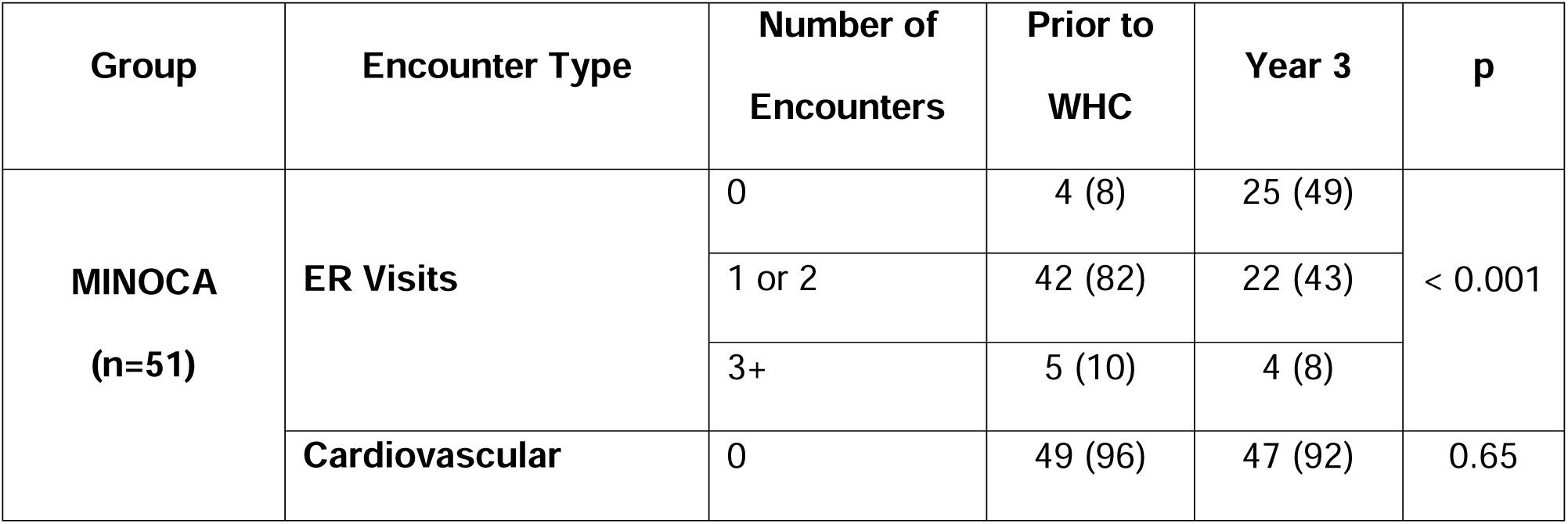
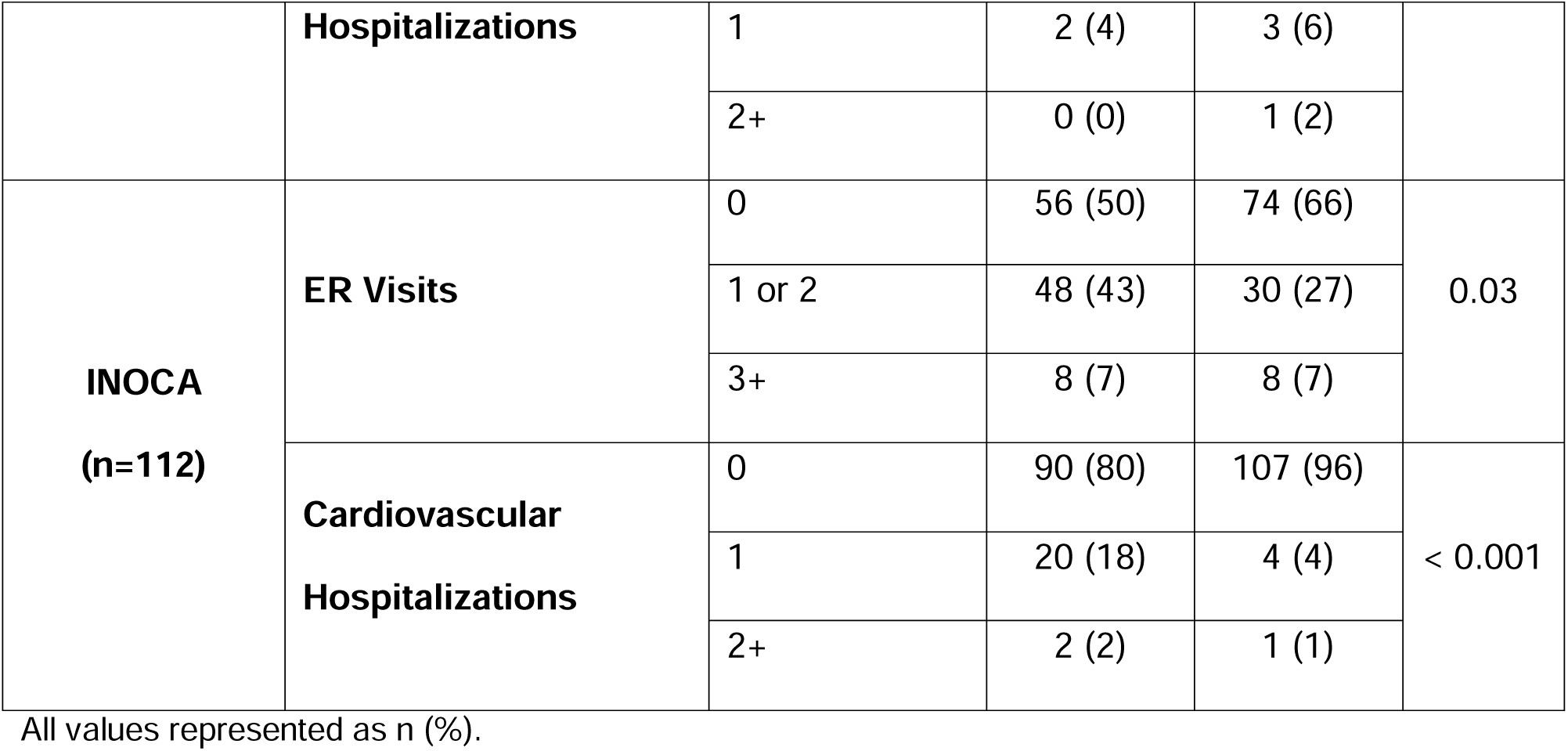
Comparing hospital encounters 3 years prior to baseline WHC appointment and 3 years following baseline.

Low rates of MACE were observed among MINOCA and INOCA patients (Table 7). All MINOCA patients’ index MI events were NSTEMIs. 49 (96%) patients had their index MI event within 3 years prior to the baseline WHC visit. In the 3 years following baseline, among MINOCA patients, one patient had an MI and subsequent PCI, one (different) patient had a TIA, and no heart failure events or deaths were observed. Among INOCA patients, 2 had a TIA event post-baseline, and one observed death was of unknown cause.

**Table 7:**
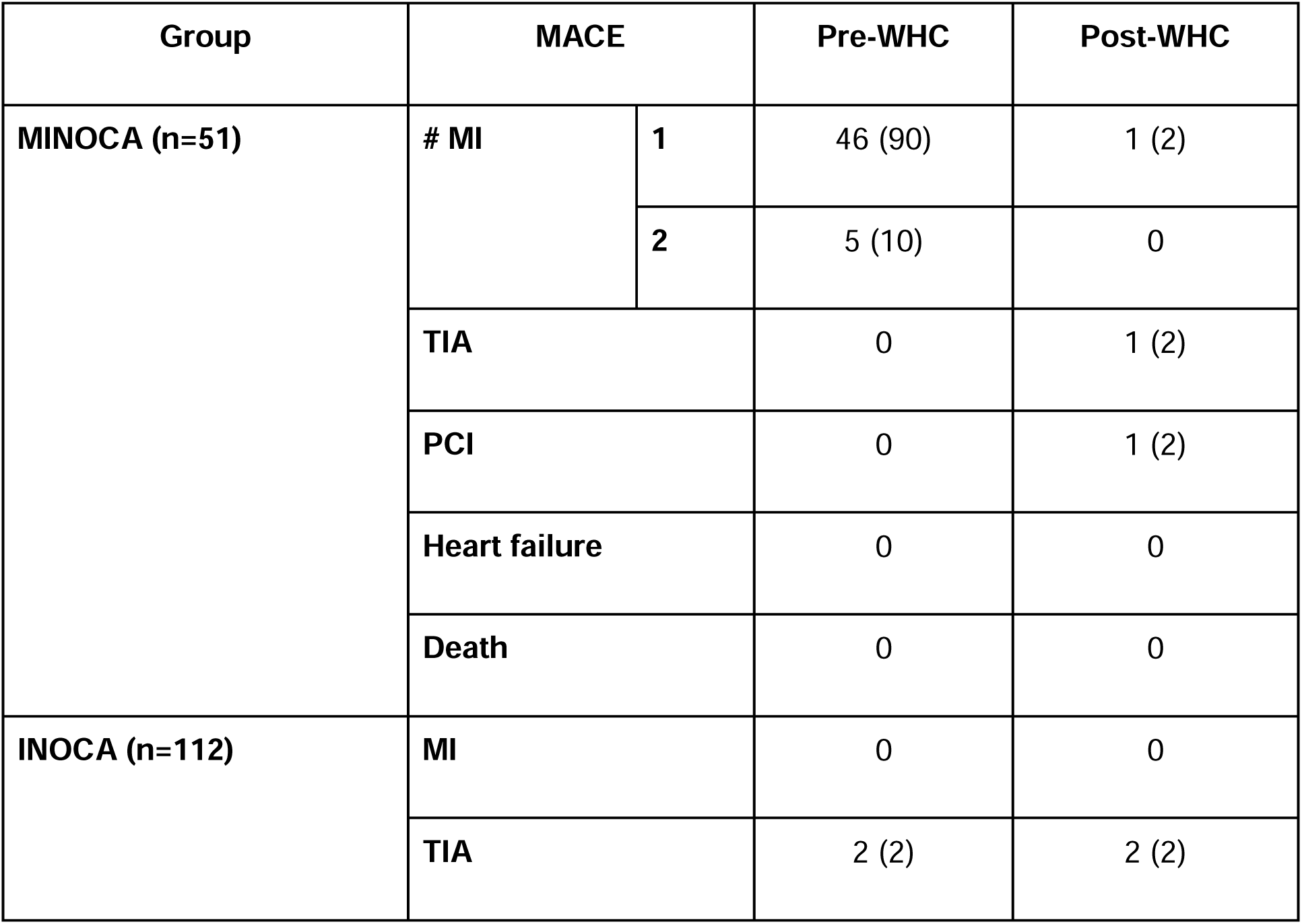
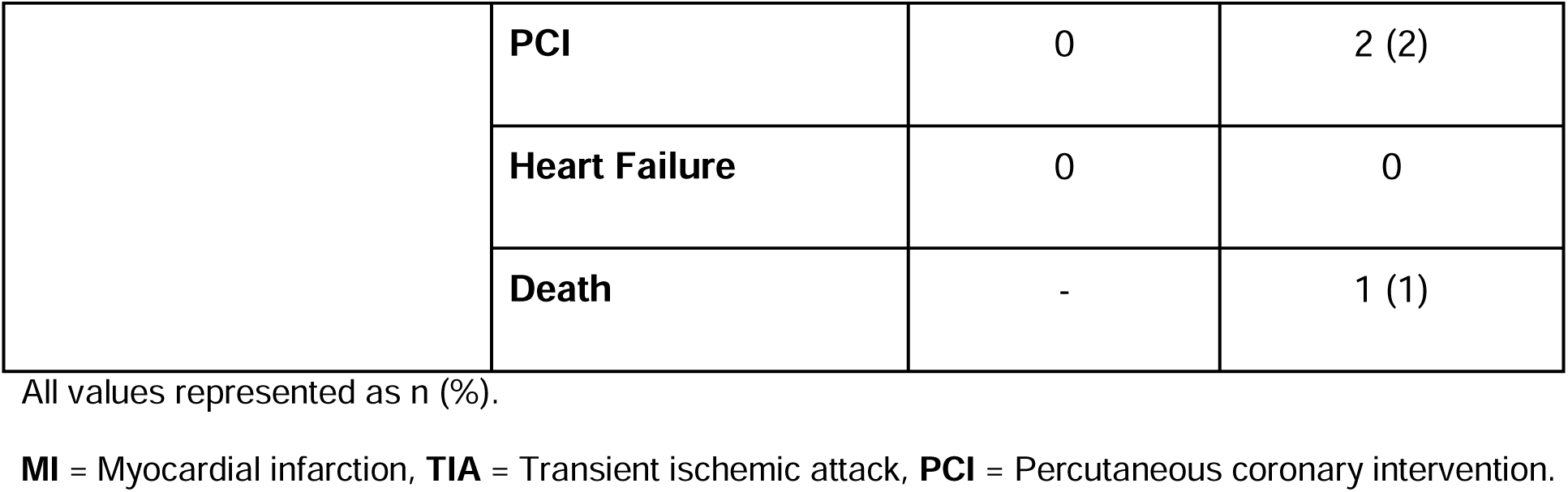
Major adverse cardiovascular event (MACE) outcomes after 3 years attendance at the WHC in (M)INOCA groups.

| **Group** | **MACE** |  | **Pre-WHC** | **Post-WHC** |
| --- | --- | --- | --- | --- |
| **MINOCA (n=51)** | # MI | 1 | 46 (90) | 1 (2) |
|  |  | 2 | 5 (10) | 0 |
|  | **TIA** |  | 0 | 1 (2) |
|  | **PCI** |  | 0 | 1 (2) |
|  | **Heart failure** |  | 0 | 0 |
|  | **Death** |  | 0 | 0 |
| **INOCA (n=112)** | **MI** |  | 0 | 0 |
|  | **TIA** |  | 2 (2) | 2 (2) |

|  |  |  |  |
| --- | --- | --- | --- |
|  | **PCI** | 0 | 2 (2) |
|  | **Heart Failure** | 0 | 0 |
|  | **Death** | - | 1 (1) |

All values represented as n (%).
**MI** = Myocardial infarction, **TIA** = Transient ischemic attack, **PCI** = Percutaneous coronary intervention.

### Therapeutic Management

Changes in the number and proportion of patients on various medical therapies are summarized in Figure 2 (Supplemental Table S1). At baseline, in quantifying the 5 most frequently used medications for both MINOCA and INOCA, MINOCA had more angiotensin-converting enzyme inhibitors/angiotensin II receptor antagonists (ACEi/ARBs) at baseline than INOCA, and INOCA had more antidepressants. Both groups shared similarly high prescriptions of acetylsalicylic acid (ASA), statins, 1,4- dihydropyridine calcium channel blockers (DHP-CCBs) and beta blockers (BBs) at baseline. At 3 years, MINOCA patients had more ASA than INOCA, and INOCA had more short-acting nitrates. Otherwise, both groups shared similarly high prescriptions of DHP-CCBs, long-acting nitrates, BBs, and statins.

**Figure 2.**
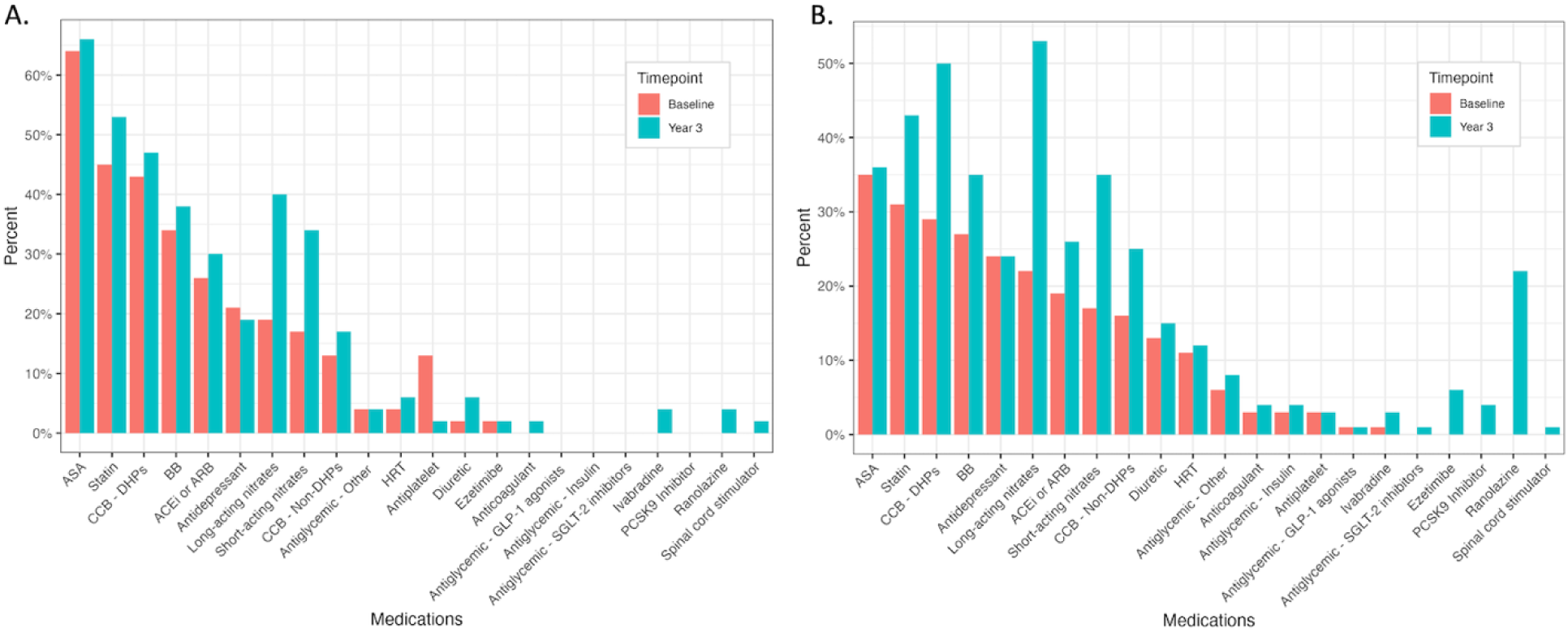
Proportions of therapies prescribed at WHC baseline and 3 years post-baseline in patients with A. MINOCA or B. INOCA. ACE. = Angiotensin-converting enzyme inhibitor, **ARB** = Angiotensin II receptor antagonist, **ASA** = Acetylsalicylic acid, **BB** = Beta blocker, **CCB** = Calcium channel blocker, **DHP** = 1,4-Dihydropyridine, **GLP-1** = Glucagon-like peptide-1, **HRT** = Hormone replacement therapy, **IDN** = Isosorbide dinitrate, **IMN** = Isosorbide mononitrate, **NSAID** = Non-steroidal anti-inflammatory drug, **PCSK9** = Proprotein convertase subtilisin/kexin type 9, **SGLT-2** = Sodium-glucose cotransporter-2

Changes in prescriptions between baseline and 3 years are summarized in Figure 3 (Supplemental Table S2). For MINOCA, the greatest increases in medications were long-acting nitrates (13, 28%), short-acting nitrates (9, 19%), BBs (8, 17%) and ASA (7, 15%). ASA, non-ASA antiplatelets and BBs demonstrated the highest rates of discontinuation (6 each, 13%). For INOCA, the greatest increases in medications were long-acting nitrates (41, 38%), DHP-CCBs (29, 27%), short-acting nitrates (27, 25%) and ranolazine (24, 22%). ASA (13, 12%) and short-acting nitrates (8, 7%) were the most frequently discontinued, followed by long-acting nitrates (7, 6%), BBs and DHP-CCBs (each 6, 6%). Medications that report high rates of both prescription and discontinuation are reflective of different patients being treated.

**Figure 3.**
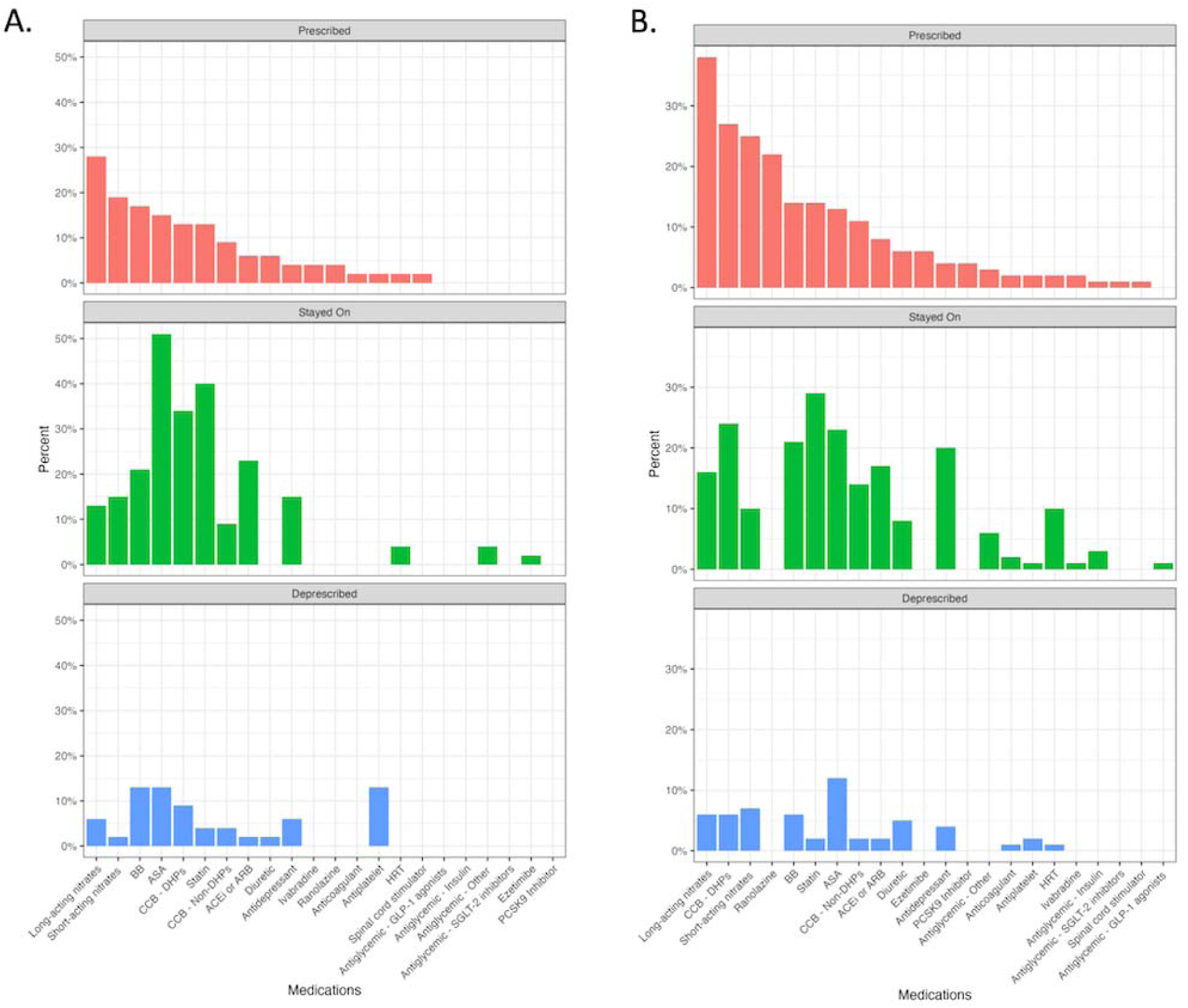
Changes in medications between WHC baseline and 3 years post-baseline for A. MINOCA and B. INOCA patients. ACE. = Angiotensin-converting enzyme inhibitor, **ARB** = Angiotensin II receptor antagonist, **ASA** = Acetylsalicylic acid, **BB** = Beta blocker, **CCB** = Calcium channel blocker, **DHP** = 1,4-Dihydropyridine, **GLP-1** = Glucagon-like peptide-1, **HRT** = Hormone replacement therapy, **IDN** = Isosorbide dinitrate, **IMN** = Isosorbide mononitrate, **NSAID** = Non-steroidal anti-inflammatory drug, **PCSK9** = Proprotein convertase subtilisin/kexin type 9, **SGLT-2** = Sodium-glucose cotransporter-2

### Patient-Reported Outcomes

Improvements in SAQ subdomain scores were noted in both MINOCA and INOCA groups between baseline and 3 years (Table 8). Among MINOCA patients, significant improvements were observed in all subdomains, except for physical limitations. In contrast, significant improvements were observed across all domains in INOCA patients. When investigating SAQ subdomains based on whether MCID were achieved, a higher relative proportion of INOCA patients were observed to have clinically meaningful improvements across all domains except treatment satisfaction compared to MINOCA patients (Figure 4, Supplemental Table S3).

**Figure 4:**
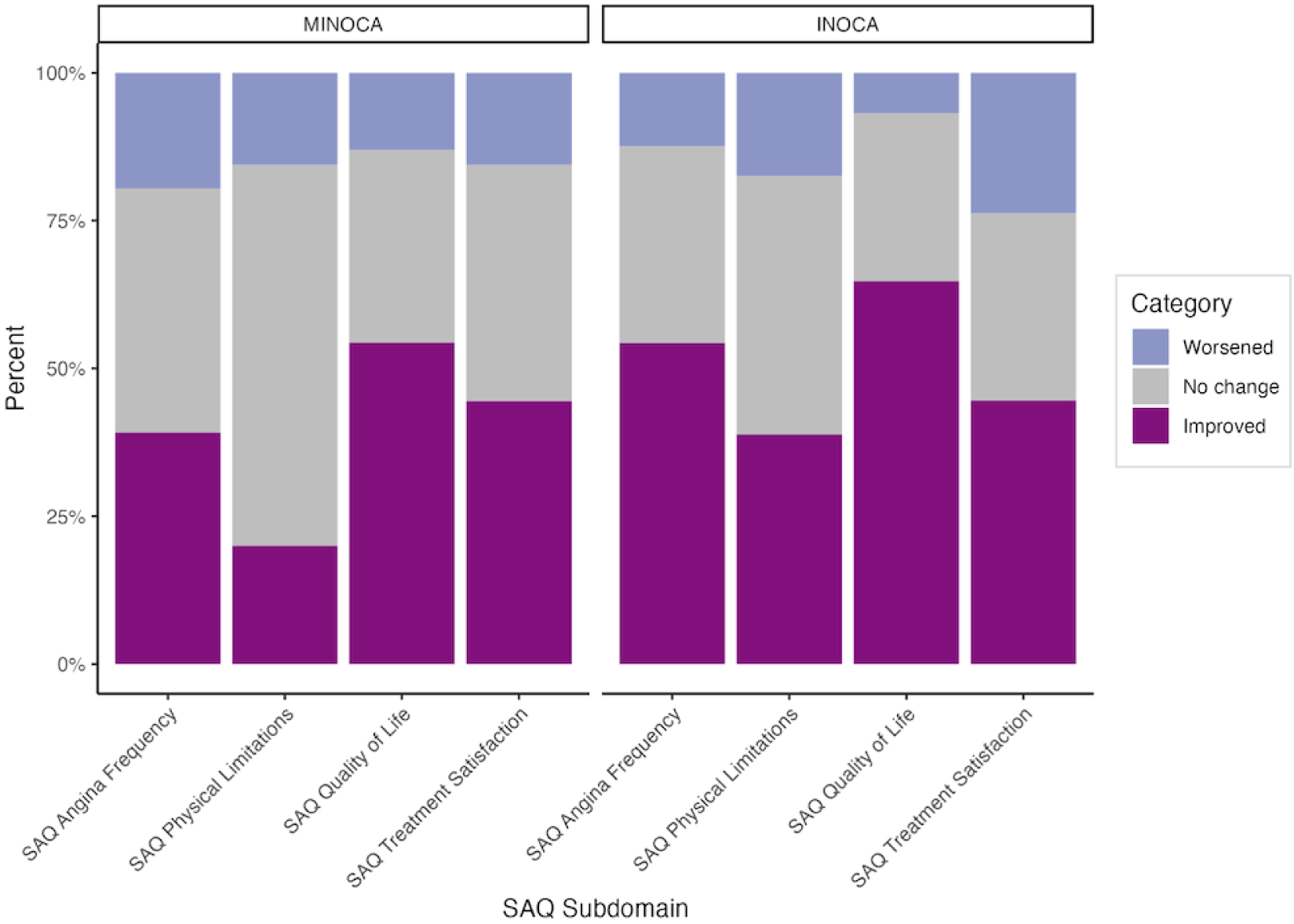
Proportion of patients who had minimal clinically important differences (MCID) in domain scores between baseline and 3 years, stratified by (M)INOCA.

**Table 8:** Comparing SAQ subdomain questionnaire scores at baseline compared to 3 years.

| **Group** | **Questionnaire** | **Baseline** | **Year 3** | **p** |
| --- | --- | --- | --- | --- |
| MINOCA<br>(n=51) | Physical Limitations | 71 (56, 86) | 71 (63, 79) | 0.47 |
|  | Angina Frequency | 60 (37, 84) | 80 (56, 100) | 0.03 |
|  | Quality of Life | 50 (34, 67) | 67 (55, 80) | < 0.001 |
|  | Treatment Satisfaction | 76 (56, 97) | 82 (70, 94) | < 0.01 |
| INOCA (n=112) | Physical Limitations | 51 (36, 66) | 60 (45, 76) | < 0.001 |
|  | Angina Frequency | 47 (31, 64) | 67 (47, 87) | < 0.001 |
|  | Quality of Life | 33 (17, 50) | 47 (53, 82) | < 0.001 |
|  | Treatment Satisfaction | 71 (59, 83) | 82 (70, 94) | 0.02 |

All values represented as median (interquartile range).

Patients with depression at baseline had significantly worse SAQ scores compared to patients without depression at baseline (all p<0.05), and at 3 years in the physical limitations (p=0.006) and angina frequency domains (p=0.03) (Figure 5, Supplemental Table S4). However, SAQ scores improved at similar rates over time between both the depression and non-depression groups. Depression at baseline was not found to be a significant modifier for any of the SAQ subdomains (Δp > 0.05).

**Figure 5.**
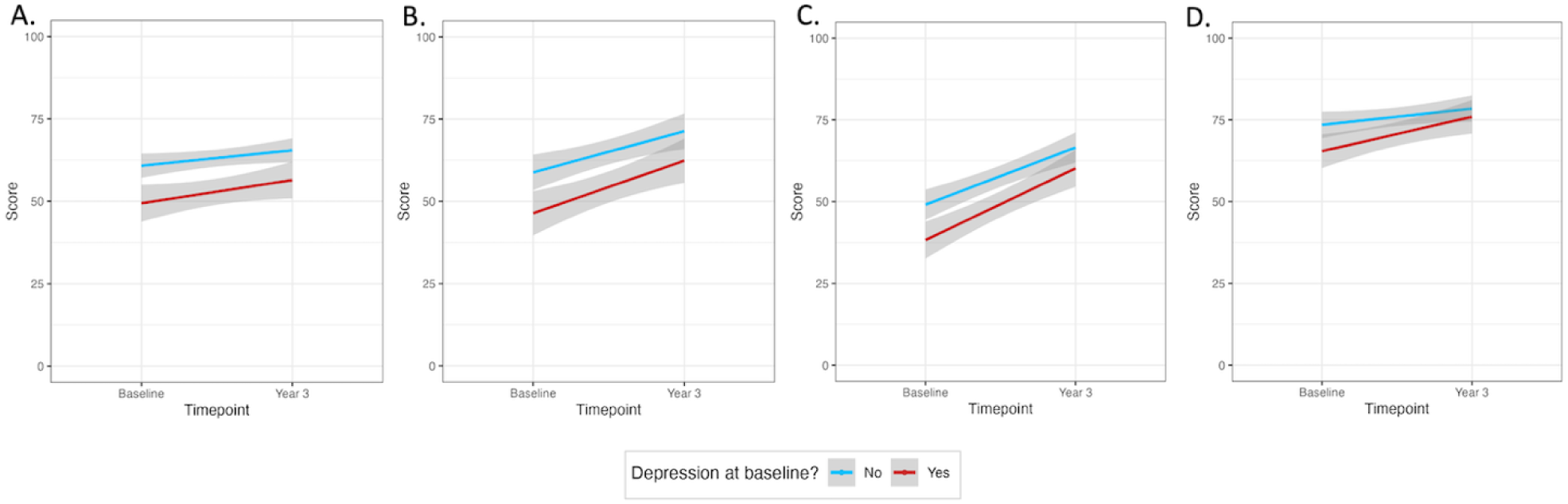
SAQ subdomain scores at baseline WHC appointment compared to 3 years late, by depression status at baseline, for: A. Physical limitations; B. Angina Frequency; C. Quality of Life; D. Treatment Satisfaction.

Patients on ranolazine at year 3 had significantly worse SAQ scores for all subdomains at baseline, and for the physical limitations and angina frequency subdomains at year 3 (Figure 6, Supplemental Table S5). SAQ scores improved at a higher rate over time in patients treated with Ranolazine at year 3 for the treatment satisfaction subdomain, suggesting that Ranolazine was an effect modifier (Δp=0.02), but it did not associate with different rates of improvement over time in the other subdomains (Δp > 0.05).

**Figure 6.**
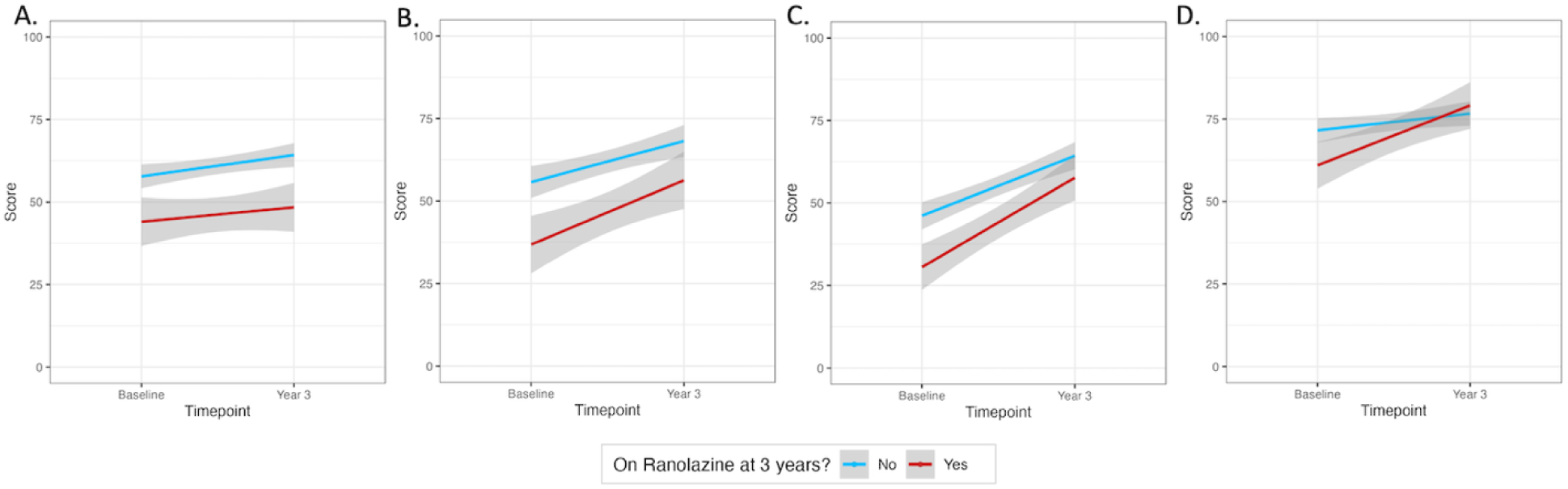
SAQ subdomain scores and standard deviation (SD) at baseline WHC appointment compared to 3 years later, by patients on ranolazine at year 3 compared to those not, for A. Physical limitations; B. Angina Frequency; C. Quality of Life; D. Treatment Satisfaction.

## Discussion

In this work, we sought to address the void of longer-term data describing (M)INOCA patients by presenting 3-year outcomes after attending a Canadian WHC. In summary, over 95% of both groups received a diagnosis, showing improved diagnostic accuracy following enrolment in a WHC. Hospital encounters, particularly emergency visits, significantly decreased; INOCA patients also experienced fewer cardiovascular hospitalizations. Medication adjustments were notable, with a reduction in non-ASA antiplatelets and a personalized approach for beta blockers. Ranolazine was frequently prescribed for INOCA patients, suggesting a challenging symptom profile in this cohort that often requires second- or third-line anti-anginal agents. Mental health at baseline was an important component towards self-reported angina, as patients with depression had worse baseline SAQ metrics; however, with WHC follow-up, they demonstrated comparable improvements to those without depression.

While 71.4% of INOCA patients and 60% of MINOCA patients had new or changed diagnoses at 1-year^14^, by 3-years of follow-up a diagnosis was achieved in 95.5% of INOCA patients and 96.1% of MINOCA patients. This improvement in diagnostic yield is suggestive of the access to testing and diagnostics through longer follow-up at the WHC including invasive coronary reactivity testing and adenosine cardiac magnetic resonance imaging.

Women with chest pain and nonobstructive coronary syndromes frequently utilize hospital resources.^22^ We demonstrated a significant reduction in emergency department presentations following enrolment and follow-up in a WHC. INOCA patients also had significantly reduced cardiovascular hospitalizations, unlike MINOCA patients, who had low baseline rates excluding their index hospitalization. MINOCA patients had more diagnostic testing performed than INOCA patients, which is likely attributed to the high-risk nature of the MI event and ability to perform more testing during their index inpatient admission.

We acknowledge the low rate of MACE endpoints, which limits our ability to detect significant differences. We suspect this may be due to the relatively short follow-up conducted, in contrast to other studies that typically report on 5- to 10-year follow-up, such as the Women’s Ischemia Syndrome (WISE) cohort, which followed female patients with predominantly non-obstructive disease. The WISE cohort reported a MACE risk of 6.7% and 12.8% after 10-years of follow-up for women with no and non-obstructive disease respectively, with earlier time points not described.^5^ Heart failure hospitalizations were also found in unexpectedly high incidence in the WISE cohort (11.2%), documented as the most common MACE following a diagnosis of CMD after being followed for 4.6-8.6 years.^24^ Importantly, the higher proportion of hypokinesis and akinesis in our MINOCA patients suggests that they may be at a higher relative risk for subsequent heart failure than INOCA patients, highlighting the importance of longer ongoing follow-up of these patients.

After 3 years of follow-up in our WHC, a number of changes were observed in medical therapies. Short- and long-acting nitrates were common medications prescribed for both MINOCA and INOCA patients and are well-recognized for their antianginal properties.^4,6^ Non-ASA antiplatelets were commonly deprescribed medications, which is supported by the lack of substantial benefit seen in the available literature particularly in INOCA.^25^ The implementation of beta blockade in patients with MINOCA remains a nuanced decision, likely explaining why this therapy was both one of the greatest prescribed and deprescribed medications over 3-years of follow-up. When Lindahl *et al.* assessed the utility of beta blockade, there was a trend towards a positive effect; however, the modality failed to reach significance.^25^ This uncertain benefit is under further review, awaiting the results of the MINOCA-BAT trial.^26^ Calcium channel blockers were one of the most prescribed medications for patients with INOCA, but not in MINOCA patients. This may be reflective of calcium channel blockers having been initiated in 42.6% of MINOCA patients prior to WHC enrolment, thus entering the WHC already on this appropriate therapy.

The 2023 JCS/CVIT/JCC guidelines on coronary vasospasm and CMD acknowledge that approximately 20% of patients may prove refractory to these therapies and warrant alternative therapeutic considerations.^27^ Ranolazine is a piperazine derivative that selectively inhibits the late sodium current, with evidence suggestive of its benefit in CMD; it can be considered as adjunctive therapy in chronic angina.^28^ Ranolazine was one of the most prescribed medications for INOCA patients from enrolment to 3-years in the WHC, with 22% initiated on the therapy as compared to 4% of MINOCA patients. This is likely multifactorial, owing to CMD being a more common etiology in INOCA patients and having a more challenging symptom profile necessitating additional line therapy.

Our study identified differences in angina through SAQ responses stratified by (M)INOCA group, depression status, and prescription of ranolazine at 3 years. INOCA patients experienced significant improvements across all SAQ subdomains. MINOCA patients did not observe a similar improvement in physical limitations; however, they significantly improved in the other subdomains of angina frequency, quality of life, and treatment satisfaction.

Patients with depression at the time of WHC enrolment had significantly worse baseline SAQ scores across all metrics, including angina frequency and quality of life. This is reflective of the literature, wherein depression has been independently associated with angina in patients without obstructive CAD.^9^ Reassuringly, depression at baseline was not a significant modifier for any subdomain over 3 years, suggesting that patients with depression improved to a similar degree to those without depression. Mental health at presentation of cardiac symptoms has important implications for the interpretation of angina severity; a women’s heart health psychiatrist is an important part of our team approach in the WHC and may help contribute to the favourable improvements seen in these patients. Acknowledging the bidirectional relationship between mental health and angina, attendance at a specialized clinic addressing both in unison also represents a plausible explanation for the improvements seen in these patients.

Improving mental health symptoms is not only an important priority for reducing symptom burden, but also reducing long-term cardiovascular outcomes. There is increasing appreciation of the interplay between mental health and cardiovascular health.^29^ From a sex-specific perspective, not only are women twice as likely as men to be diagnosed with depression, the INTERHEART study found psychosocial factors increase the odds of MI by 3.5-fold in women as compared to 2.6-fold in men.^30^

In this study, patients prescribed ranolazine by year 3 had significantly worse SAQ scores for all subdomains at the time of WHC enrolment. These patients subsequently improved by a similar degree across time points to patients not on ranolazine, except with regards to medication satisfaction, where ranolazine was a significant modifier towards greater improvement over time. This medication, which is new to Canada, is an important adjunctive agent and should be considered early in treatment.

## Limitations

WHCs are relatively novel entities, with our centre being one of 6 in Canada and the only center in British Columbia. As such, and by nature of our single-center data collection without a control group, this inherently brings with it limitations. Our patient population is predominantly Caucasian and other ethnicities are underrepresented. Further, due to referral of SCAD patients to a dedicated SCAD clinic rather than our WHC, this patient population is under-represented in this dataset compared to other studies of (M)INOCA patients. Therefore, the generalizability of our results to other WHCs with more representation of SCAD and other ethnicities is unclear. Further, measures of gender identity were not captured. It is unclear the extent to which our findings extend to male patients with nonobstructive vasomotor etiologies.

## Conclusion

This study addresses a gap in longer-term data for (M)INOCA patients by presenting 3- year outcomes after specialized care in a Canadian WHC. The enhanced diagnostic clarity and the differentiation of symptom characteristics between these patient groups underscore the importance of specialized continuity of care. These findings suggest that ongoing care through a WHC, including attention to mental health and tailored medication strategies, is crucial for achieving optimal outcomes in (M)INOCA patients. Acknowledging the practical implications of 6 available WHCs across Canada, knowledge dissemination is imperative in bridging the gaps for patients awaiting specialized access. When appropriate, virtual healthcare can be beneficial. Additionally, clinical trial and observational data to evaluate the efficacy of different medication regimens on MACE outcomes is warranted across larger cohorts representing more diverse gender and ethnic identities. Future directions entail establishment of a control group in facilitating robust comparative assessments of long-term data.

## Data Availability

Data produced in this study are available upon reasonable request to the authors.

## Acknowledgements

The authors would like to thank the patients attending the WHC for their participation in this research; statistician May Lee for her valuable consultation in statistical analysis; and Lily Cai, Andrew Starovoytov, Chenille Wong and Sasha Voznyuk in data collection.

The authors confirm that patient consent forms have been obtained for the WHC Registry through the University of British Columbia clinical research ethics board application #H13-03322. Secondary consent was not required for this article as aggregate deidentified data were used for the analyses as within scope of the WHC Registry protocol.

