## Supplemental Material for "Unveiling the Longitudinal Journey: Three-Year Follow-up of Women with MINOCA and INOCA in a Specialized Heart Centre"

**Supplemental Methods
Definitions of Baseline Characteristics**

All variables are per diagnosis at baseline, unless indicated otherwise (e.g. “history of…”).

| **Variable** | **Definition** |
| --- | --- |
| ***Demographics*** | |
| **Age** | Patient’s age at first clinic visit, calculated by subtracting the year of their first WHC visits by their year of birth. |
| **Race** | Self-reported with options: African-Canadian, Caucasian, East Asian, First Nation, South Asian, Other. “Other” includes mixed ancestry. As the large majority of this cohort identified as solely Caucasian, this variable was made into a binary. |
| **Partnered status** | Self-reported marital status, categorized into a binary wherein “partnered” refers to common law or married, and not partnered refers to single, widowed or divorced. |
| **Education** | Self-reported with options: High school or less, some post-secondary (any amount of college or university up to and including a Bachelor’s degree), and post-secondary above a Bachelor’s (e.g., law degree, Master’s, doctorate, PharmD, MD, etc.). |
| **Working status** | Self-reported and categorized as employed, not employed” (including if retired), or on medical leave/disability. |
| ***Clinical*** | |
| **Family history of premature cardiovascular disease** | Self-reported as any first-degree relative (parent, sibling, child) having had a cardiovascular diagnosis under 65 years old for women or under 55 years old for men. |
| **Dyslipidemia** | Defined in consults as total cholesterol ≥ 240 mg/dL [6.2 mmol/L], low density lipoprotein cholesterol ≥ 160 mg/dL [4.1 mmol/L], high-density lipoprotein cholesterol ≤ 40 mg/dL [1.0 mmol/L] and triglycerides ≥ 200 mg/dL [2.3 mmol/L]^1^. |
| **Diabetes** | Defined in consults as fasting plasma glucose ≥ 126 mg/dL (7 mmol/L) in at least 2 baseline measurements and a hemoglobinA1c ≥ 6.5%^2^. |
| **Hypertension** | Defined in consults as systolic/diastolic blood pressure ≥140/90mmHg per the Canadian hypertension guidelines^3^. |
| **Obesity** | Body mass index (BMI) of ≥30 kg/m^2^, calculated from the most recent height and weight measurements in the patient’s health records^4^. |
| **Smoking history** | Self-reported as current, former, or never smoker. |
| **Chronic kidney dysfunction** | Indicated in consults as defined by abnormalities in either kidney structure or function for >3 months including an estimated glomerular filtration rate (eGFR) of under 60 mL/min/1.73 m^2^, albuminuria >3 mg/mmol by albumin to creatinine ratio, urine sediment abnormalities, kidney transplant history, or structural abnormalities detected by imaging^5^. |
| **History of** **arrhythmia** | Indicated in past or current physician consults such as atrial fibrillation, sick sinus syndrome, ventricular tachycardia or ventricular fibrillation. |
| **History of prior transient ischemic attack (TIA)** | As diagnosed by a neurologist or other attending specialist and is defined as a temporary episode of neurological dysfunction caused by a brief disruption cerebral perfusion^6^. |
| **Thyroid dysfunction** | A thyroid disorder of hypothyroidism or hyperthyroidism indicated in consults as defined by triiodothyronine (T3), thyroxine (T4) and thyroid stimulating hormone (TSH) levels under or over clinical thresholds^7,8^. Thyroid dysfunction due to an autoimmune cause (e.g., Grave’s, Hashimoto’s), infectious (e.g., thyroiditis) or surgical causes requiring supplemental thyroid medications (e.g., thyroidectomy) are included. |
| **History of migraines** | Indicated in past or current physician consults, and/or self-report, defined as a type of headache characterized by recurrent attacks of moderate to severe unilateral pulsating pain^9^. |
| **History of Raynaud’s syndrome** | Indicated in past or current physician consults, and/or self-report, defined as episodic vasospasm to the distal extremities, usually triggered by exposure to cold temperatures^10^. |
| **History of chronic obstructive pulmonary disease (COPD)** | Indicated in past or current physician consults, defined as a pulmonary disease characterized by chronic respiratory symptoms due to abnormalities in the airways and/or alveoli, leading to chronic progressive airflow obstruction^11^. |
| **History of any autoimmune disease** | Indicated in past or current physician consults. |
| **History of clinical anxiety** | Indicated in past or current physician consults, and encompasses anxiety-related diagnoses of post-traumatic stress disorder, panic disorder, and phobias^12^. |
| ***Female-specific*** | |
| **Menopause status** | Indicated in physician consults and confirmed by self-report as pre-, peri-, post-, or surgical due to a hysterectomy. Surgical and post-menopausal status was defined as not having had a menstrual period for over 6 months^13^. Peri-menopausal status is defined as the transitional phase preceding menopause, characterized by hormonal fluctuations, irregular menstrual cycles, and the onset of vasomotor symptoms, marking the end of reproductive capacity^13^. |
| **Number of children born** | Self-reported as number of births in the patient’s lifetime. The number of children in multiparous pregnancies are included in this count. |
| **Preeclampsia or hypertension in pregnancy** | Indicated in past or current physician consults, and/or self-report, and is defined as gestational systolic/diastolic blood pressure ≥140/90 mmHg (with proteinuria, for preeclampsia)^14^. |
| **Polycystic ovarian syndrome (PCOS)** | Indicated in past or current physician consults, and/or self-report as defined by two of: oligo- and/or anovulation; clinical and/or biochemical signs of hyperandrogenism; imaging evidence of polycystic ovaries^15^. |

**Definitions of Vasomotor Diagnoses**

Regarding specific etiologies, for both CMD and coronary vasospasm, we used the Coronary Vasomotion Disorder International Study Group (COVADIS) definition of probable and definite^16^. In brief, definitive CMD is diagnosed when all 4 the following criteria were met: (1) symptoms of myocardial ischemia, (2) Absence of CAD (<50% stenosis in any epicardial artery), (3) objective evidence of ischemia (on exercise stress testing or stress imaging), and (4) evidence of impaired coronary microvascular function on coronary flow reserve invasive testing by coronary flow reserve (CFR) or index of microcirculatory resistance (IMR)^16^. Probable CMD includes the first three criteria but not the fourth^16^. Regarding coronary vasospasm, the COVADIS group developed three criteria for diagnosis including (1) nitrate responsive angina (2) transient ischemic ECG changes including >1mm of ST depression or elevation at the time of symptoms and (3) transient >90% constriction of an epicardial artery during coronary angiography either spontaneously or in response to a provocative stimulus such as acetylcholine or ergonovine. Definite vasospasm is defined as the first criteria plus either the second or the third criteria. Probable angina is defined as the first criteria without the second or third^17^.

**Supplemental Methods References**
^1^ National Cholesterol Education Program (NCEP) Expert Panel on Detection, Evaluation, and Treatment of High Blood Cholesterol in Adults (Adult Treatment Panel III). Third Report of the National Cholesterol Education Program (NCEP) Expert Panel on Detection, Evaluation, and Treatment of High Blood Cholesterol in Adults (Adult Treatment Panel III) final report. Circulation. 2002;106(25):3143-421.

^2^ Diabetes Canada Clinical Practice Guidelines Expert Committee, Punthakee Z, Goldenberg R, Katz P. Definition, Classification and Diagnosis of Diabetes, Prediabetes and Metabolic Syndrome. Can J Diabetes. 2018;42 Suppl 1:S10-S15.

^3^ Rabi DM, McBrien KA, Sapir-Pichhadze R, et al. Hypertension Canada's 2020 Comprehensive Guidelines for the Prevention, Diagnosis, Risk Assessment, and Treatment of Hypertension in Adults and Children. Can J Cardiol. 2020;36(5):596-624.

^4^ Government of Canada. Canadian Guidelines for Body Weight Classification in Adults - Quick Reference Tool for Professionals. Available at: [https://www.canada.ca/en/health-canada/services/food-nutrition/healthy-eating/healthy-weights/canadian-guidelines-body-weight-classification-adults/quick-reference-tool-professionals.html.](https://www.canada.ca/en/health-canada/services/food-nutrition/healthy-eating/healthy-weights/canadian-guidelines-body-weight-classification-adults/quick-reference-tool-professionals.html.%202016) 2016. [Accessed on February 4, 2024]

^5^ Stevens PE, Levin A. Kidney Disease: Improving Global Outcomes Chronic Kidney Disease Guideline Development Work Group Members. Evaluation and management of chronic kidney disease: synopsis of the kidney disease: improving global outcomes 2012 clinical practice guideline. Ann Intern Med. 2013;158(11):825-30.

^6^ Boulanger JM, Lindsay MP, Gubitz G, et al. Canadian Stroke Best Practice Recommendations for Acute Stroke Management: Prehospital, Emergency Department, and Acute Inpatient Stroke Care. Int J Stroke. 2018;13(9):949-984.

^7^ Jonklaas J, Bianco AC, Bauer AJ, et al. American Thyroid Association Task Force on Thyroid Hormone Replacement. Guidelines for the treatment of hypothyroidism: prepared by the American thyroid association task force on thyroid hormone replacement. Thyroid. 2014;24(12):1670-751.

^8^ Ross DS, Burch HB, Cooper DS, et al. 2016 American Thyroid Association Guidelines for Diagnosis and Management of Hyperthyroidism and Other Causes of Thyrotoxicosis. Thyroid. 2016;26(10):1343-1421.

^9^ National Institutes of Health: Neurological Disorders and Stroke. Migraine. Available at: <https://www.ninds.nih.gov/health-information/disorders/migraine>. 2023. [Accessed on February 4, 2024]

^10^ Block JA, Sequeira W. Raynaud's phenomenon. Lancet. 2001;357(9273):2042-8.

^11^ Global Initiative for Chronic Obstructive Lung Disease. 2024 GOLD Report. Available at: <https://goldcopd.org/2024-gold-report/>. 2024. [Accessed on February 4, 2024]

^12^ Katzman MA, Bleau P, Blier P,et al. Canadian clinical practice guidelines for the management of anxiety, posttraumatic stress and obsessive-compulsive disorders. BMC Psychiatry. 2014;14 Suppl 1(Suppl 1):S1.

^13^ Yuksel N, Evaniuk D, Huang L, et al. Guideline No. 422a: Menopause: Vasomotor Symptoms, Prescription Therapeutic Agents, Complementary and Alternative Medicine, Nutrition, and Lifestyle. J Obstet Gynaecol Can. 2021;43(10):1188-1204.e1.

^14^ Gestational Hypertension and Preeclampsia: ACOG Practice Bulletin, Number 222. Obstet Gynecol. 2020;135(6):e237-e260.

^15^ Legro RS, Arslanian SA, Ehrmann DA, Hoeger KM, Murad MH, Pasquali R, Welt CK; Endocrine Society. Diagnosis and treatment of polycystic ovary syndrome: an Endocrine Society clinical practice guideline. J Clin Endocrinol Metab. 2013;98(12):4565-92.

^16^ Ong P, Camici PG, Beltrame JF, et al. Coronary Vasomotion Disorders International Study Group (COVADIS). International standardization of diagnostic criteria for microvascular angina. Int J Cardiol. 2018;250:16-20.

^17^ Beltrame JF, Crea F, Kaski JC, et al. Coronary Vasomotion Disorders International Study Group (COVADIS). International standardization of diagnostic criteria for vasospastic angina. Eur Heart J. 2017;38(33):2565-2568.

**Supplemental Tables**

**Supplemental Table S1. Counts and proportions of medications and therapies in patients with MINOCA and INOCA by timepoint.**

| **Category** | **Class** | **Medication** | **MINOCA  (n=47)** | | **INOCA (n=109)** | |
| --- | --- | --- | --- | --- | --- | --- |
|  |  |  | **Baseline** | **3yr** | **Baseline** | **3yr** |
| Antiplatelets/ Anticoagulants | Antiplatelet | ASA | 30 (64) | 31 (66) | 38 (35) | 39 (36) |
|  |  | Any non-ASA | 6 (13) | 3 (3) | 3 (3) | 3 (3) |
|  | Anticoagulant | Any | 0 (0) | 1 (2) | 3 (3) | 4 (4) |
| Anti-Anginal | BB | Any | 16 (34) | 18 (38) | 29 (27) | 38 (35) |
|  | CCBs | DHPs (amlodipine, nifedipine) | 20 (43) | 22 (47) | 32 (29) | 55 (50) |
|  |  | Non-DHPs (diltiazem, verapamil) | 6 (13) | 8 (17) | 17 (16) | 27 (25) |
|  | Nitrates | Short-acting nitrates (spray) | 8 (17) | 16 (34) | 19 (17) | 38 (35) |
|  |  | Long-acting nitrates (patch, IMN or IDN) | 9 (19) | 19 (40) | 24 (22) | 58 (53) |
|  | Other | Ivabradine | 0 (0) | 2 (4) | 1 (1) | 3 (3) |
|  |  | Ranolazine | 0 (0) | 2 (4) | 0 (0) | 24 (22) |
| Cholesterol-Lowering | Statin | Any | 21 (45) | 25 (53) | 34 (31) | 47 (43) |
|  | Other | Ezetimibe | 1 (2) | 1 (2) | 0 (0) | 6 (6) |
|  |  | PCSK9 Inhibitor | 0 (0) | 0 (0) | 0 (0) | 4 (4) |
| Other Therapies | ACE/ARB | Any | 12 (26) | 14 (30) | 21 (19) | 28 (26) |
|  | Diuretic | Any | 1 (2) | 3 (6) | 14 (13) | 16 (15) |
|  | Antiglycemic | Insulin | 0 (0) | 0 (0) | 3 (3) | 4 (4) |
|  |  | GLP-1 agonists (eg. semaglutide) | 0 (0) | 0 (0) | 1 (1) | 1 (1) |
|  |  | SGLT-2 inhibitors (eg. empagliflozin) | 0 (0) | 0 (0) | 0 (0) | 1 (1) |
|  |  | Other (incl. metformin) | 2 (4) | 2 (4) | 6 (6) | 9 (8) |
|  | Antidepressant | Any | 10 (21) | 9 (19) | 26 (24) | 26 (24) |
|  | HRT | Any | 2 (4) | 3 (6) | 12 (11) | 13 (12) |
|  |  | Spinal cord stimulator | 0 (0) | 1 (2) | 0 (0) | 1 (1) |

All values presented as n (%).
P-values denoted by hyphen (-) reflect too few n in at least one cell (n<5) in the group (MINOCA or INOCA) to interpret statistical significance.

**ACE** = Angiotensin-converting enzyme inhibitor, **ARB** = Angiotensin II receptor antagonist, **ASA** = Acetylsalicylic acid, **BB** = Beta blocker, **CCB** = Calcium channel blocker, **DHP** = 1,4-Dihydropyridine, **GLP-1** = Glucagon-like peptide-1, **HRT** = Hormone replacement therapy, **IDN** = Isosorbide dinitrate, **IMN** = Isosorbide mononitrate, **PCSK9** = Proprotein convertase subtilisin/kexin type 9, **SGLT-2** = Sodium-glucose cotransporter-2

**Supplemental Table S2. Changes in therapies between WHC baseline and 3 years post-baseline.**

| **Category** | **Class** | **Medication** | **MINOCA (n=47)** | | | | **INOCA (n=109)** | | | |
| --- | --- | --- | --- | --- | --- | --- | --- | --- | --- | --- |
|  |  |  | **On pre-WHC and stayed on** | **Prescribed in WHC** | **Taken off in WHC** | **Never on** | **On pre-WHC and stayed on** | **Prescribed in WHC** | **Taken off in WHC** | **Never on** |
| Antiplatelets/ Anticoagulants | Antiplatelet | ASA | 24 (51) | 7 (15) | 6 (13) | 10 (21) | 25 (23) | 14 (13) | 13 (12) | 57 (52) |
|  |  | Any non-ASA | 0 (0) | 1 (2) | 6 (13) | 40 (85) | 1 (1) | 2 (2) | 2 (2) | 104 (95) |
|  | Anticoagulant | Any | 0 (0) | 1 (2) | 0 (0) | 46 (98) | 2 (2) | 2 (2) | 1 (1) | 104 (95) |
| Anti-Anginal | BB | Any | 10 (21) | 8 (17) | 6 (13) | 23 (49) | 23 (21) | 15 (14) | 6 (6) | 65 (60) |
|  | CCBs | DHPs (amlodipine, nifedipine) | 16 (34) | 6 (13) | 4 (9) | 21 (45) | 26 (24) | 29 (27) | 6 (6) | 48 (44) |
|  |  | Non-DHPs (diltiazem, verapamil) | 4 (9) | 4 (9) | 2 (4) | 37 (79) | 15 (14) | 12 (11) | 2 (2) | 80 (73) |
|  | Nitrates | Short-acting nitrates (spray) | 7 (15) | 9 (19) | 1 (2) | 30 (64) | 11 (10) | 27 (25) | 8 (7) | 63 (58) |
|  |  | Long-acting nitrates (patch, IMN or IDN) | 6 (13) | 13 (28) | 3 (6) | 25 (53) | 17 (16) | 41 (38) | 7 (6) | 44 (40) |
|  | Other | Ivabradine | 0 (0) | 2 (4) | 0 (0) | 45 (96) | 1 (1) | 2 (2) | 0 (0) | 106 (97) |
|  |  | Ranolazine | 0 (0) | 2 (4) | 0 (0) | 45 (96) | 0 (0) | 24 (22) | 0 (0) | 85 (78) |
| Cholesterol-Lowering | Statin | Any | 19 (40) | 6 (13) | 2 (4) | 20 (43) | 32 (29) | 15 (14) | 2 (2) | 60 (55) |
|  | Other | Ezetimibe | 1 (2) | 0 (0) | 0 (0) | 46 (98) | 0 (0) | 6 (6) | 0 (0) | 103 (94) |
|  |  | PCSK9 Inhibitor | 0 (0) | 0 (0) | 0 (0) | 47 (100) | 0 (0) | 4 (4) | 0 (0) | 105 (96) |
| Other | ACE/ARB | Any | 11 (23) | 3 (6) | 1 (2) | 32 (68) | 19 (17) | 9 (8) | 2 (2) | 79 (72) |
|  | Diuretic | Any | 0 (0) | 3 (6) | 1 (2) | 43 (91) | 9 (8) | 7 (6) | 5 (5) | 88 (81) |
|  | Antiglycemic | Insulin | 0 (0) | 0 (0) | 0 (0) | 47 (100) | 3 (3) | 1 (1) | 0 (0) | 105 (96) |
|  |  | GLP-1 agonists (eg. semaglutide) | 0 (0) | 0 (0) | 0 (0) | 47 (100) | 1 (1) | 0 (0) | 0 (0) | 108 (99) |
|  |  | SGLT-2 inhibitors (eg. empagliflozin) | 0 (0) | 0 (0) | 0 (0) | 47 (100) | 0 (0) | 1 (1) | 0 (0) | 108 (99) |
|  |  | Other (incl. metformin) | 2 (4) | 0 (0) | 0 (0) | 45 (96) | 6 (6) | 3 (3) | 0 (0) | 100 (92) |
|  | Antidepressant | Any | 7 (15) | 2 (4) | 3 (6) | 35 (74) | 22 (20) | 4 (4) | 4 (4) | 79 (72) |
|  | HRT | Any | 2 (4) | 1 (2) | 0 (0) | 44 (94) | 11 (10) | 2 (2) | 1 (1) | 95 (87) |
|  | Spinal cord stimulator |  | 0 (0) | 1 (2) | 0 (0) | 46 (98) | 0 (0) | 1 (1) | 0 (0) | 108 (99) |

All values presented as n (%).

**ACE** = Angiotensin-converting enzyme inhibitor, **ARB** = Angiotensin II receptor antagonist, **ASA** = Acetylsalicylic acid, **BB** = Beta blocker, **CCB** = Calcium channel blocker, **DHP** = 1,4-Dihydropyridine, **GLP-1** = Glucagon-like peptide-1, **HRT** = Hormone replacement therapy, **IDN** = Isosorbide dinitrate, **IMN** = Isosorbide mononitrate, **PCSK9** = Proprotein convertase subtilisin/kexin type 9, **SGLT-2** = Sodium-glucose cotransporter-2

**Supplemental Table S3. Comparing counts of participants that had changes in SAQ subdomain questionnaire scores from baseline to 3 years later that were above the questionnaire-specific minimal clinically important difference (MCID) range (“improved”), within the MCID range (“no change”) or below the MCID range (“worsened”).**

| **Group** | **MCID category** | **Physical Limitations** | **Angina Frequency** | **Quality of Life** | **Treatment Satisfaction** |
| --- | --- | --- | --- | --- | --- |
| MINOCA (n=51) | Improved | 9 (18) | 18 (35) | 25 (49) | 20 (39) |
|  | No change | 29 (57) | 19 (37) | 15 (29) | 18 (35) |
|  | Worsened | 7 (14) | 9 (18) | 6 (12) | 7 (14) |
|  | *Missing* | *6 (12)* | *5 (10)* | *5 (10)* | *6 (12)* |
| INOCA (n=112) | Improved | 40 (36) | 57 (51) | 66 (59) | 45 (40) |
|  | No change | 45 (40) | 35 (31) | 29 (26) | 32 (29) |
|  | Worsened | 18 (16) | 13 (12) | 7 (6) | 24 (21) |
|  | *Missing* | *9 (8)* | *7 (6)* | *10 (9)* | *11 (10)* |

All values represented as n (%).

**Supplemental Table S4. Mean questionnaire scores for SAQ subdomains within timepoints (baseline, year 3) between group status (depression at baseline or not).** p describes if there are  significant differences between means of each group (depression at baseline or not) within each timepoint. Δp describes if there are significant differences between the slopes between each group across timepoints.

| **SAQ Subdomain** | **Baseline mean (SD)** | | | **Year 3 mean (SD)** | | | **Δ scores (Y3-Baseline)** | | |
| --- | --- | --- | --- | --- | --- | --- | --- | --- | --- |
|  | **Depression at baseline (n=71)** | **No depression at baseline (n=92)** | **p** | **Depression at baseline (n=71)** | **No depression at baseline (n=92)** | **p** | **Depression at baseline (n=71)** | **No depression at baseline (n=92)** | **Δp** |
| **Physical limitations** | 50 (22) | 61 (18) | < 0.001 | 56 (23) | 66 (16) | 0.006 | 7 (18) | 5 (17) | 0.42 |
| **Angina frequency** | 47 (27) | 59 (26) | 0.006 | 62 (28) | 72 (24) | 0.03 | 16 (24) | 13 (25) | 0.39 |
| **Quality of Life** | 38 (24) | 48 (22) | 0.006 | 60 (22) | 67 (20) | 0.06 | 22 (21) | 17 (26) | 0.25 |
| **Treatment Satisfaction** | 66 (22) | 73 (18) | 0.02 | 76 (20) | 78 (18) | 0.43 | 11 (26) | 5 (23) | 0.19 |

**Supplemental Table S5. Mean questionnaire scores and standard deviation (SD) for SAQ subdomains and summary scores within timepoints (baseline, year 3) between group status (ranolazine at year 3 or not).** p describes if there are  significant differences between means of each group (ranolazine at year 3 or not) within each timepoint. Δp describes if there are significant differences between the slopes between each group across timepoints.

| **SAQ Subdomain** | **Baseline mean (SD)** | | | **Year 3 mean (SD)** | | | **Δ scores (Y3-Baseline)** | |  |
| --- | --- | --- | --- | --- | --- | --- | --- | --- | --- |
|  | **Ranolazine at year 3 (n=26)** | **No ranolazine at year 3 (n=137)** | **p** | **Ranolazine at year 3 (n=26)** | **No ranolazine at year 3 (n=137)** | **p** | **Ranolazine at year 3 (n=26)** | **No ranolazine at year 3 (n=137)** | **Δp** |
| Physical limitations | 44 (17) | 58 (21) | < 0.001 | 48 (19) | 64 (19) | < 0.001 | 4 (16) | 6 (18) | 0.56 |
| Angina frequency | 36 (22) | 57 (27) | < 0.001 | 56 (22) | 70 (27) | 0.01 | 19 (23) | 12 (25) | 0.18 |
| Quality of Life | 30 (16) | 46 (24) | < 0.001 | 58 (18) | 65 (22) | 0.08 | 27 (23) | 18 (25) | 0.09 |
| Treatment Satisfaction | 61 (20) | 72 (20) | 0.01 | 79 (15) | 77 (19) | 0.54 | 18 (24) | 5 (25) | 0.02 |
